# High Sensitivity and NPV for BinaxNOW Rapid Antigen Test in Children at a Mass Testing Site During Prevalent Delta Variant Period

**DOI:** 10.1101/2022.01.05.22268788

**Authors:** Kristie J. Sun, Mary Jane E. Vaeth, Matthew Robinson, Maryam Elhabashy, Ishaan Gupta, Sophia Purekal, E. Adrianne Hammershaimb, Ria Peralta, Asia Mitchell, Maisha Foyez, J. Kristie Johnson, James R Ficke, Yukari C. Manabe, James D. Campbell, Charles W. Callahan, Charles F. Locke, Melinda Kantsiper, CONQUER COVID Consortium, Zishan K. Siddiqui

## Abstract

SARS-CoV-2 continues to develop new, increasingly infectious variants including delta and omicron. We evaluated the efficacy of the Abbott BinaxNOW Rapid Antigen Test against Reverse Transcription Polymerase Chain Reaction (“RT-PCR”) in 1054 pediatric participants presenting to a high-volume Coronavirus Disease 2019 (COVID-19) testing site while the delta variant was predominant. Participants were grouped by COVID-19 exposure and symptom status. RT-PCR demonstrated an overall prevalence of 5.2%. For all participants, sensitivity of the BinaxNOW was 92.7% (95% CI 82.4%-98.0%) and specificity was 98.0% (95% CI 97.0%-98.8%). For symptomatic participants, positive predictive value (PPV) was 72.7% (95% CI 54.5%-86.7%) and negative predictive value (NPV) was 99.2% (95% CI 98.2%-100%). Among asymptomatic participants, PPV was 71.4% (95% CI 53.7%-85.4%) and NPV was 99.7% (95% CI 99.0%-100%). Our reported sensitivity and NPV are higher than other pediatric studies, potentially because of higher viral load from the delta variant, but specificity and PPV are lower.

**Importance:** The BinaxNOW rapid antigen COVID-19 test had a sensitivity of nearly 92% in both symptomatic and asymptomatic children when performed at a high-throughput setting during the more transmissible delta variant dominant period. The test may play an invaluable role in asymptomatic screening and keeping children safe in school.

## INTRODUCTION

Many U.S. children remain susceptible to SARS-CoV-2 infection. 20 million children under 5 have no option for vaccination.^1^ Among the 28 million eligible children aged 5-11, 18% have received at least one dose of the Coronavirus Disease 2019 (COVID-19) vaccine as of December 8, 2021.^2^ Similarly, only 51% of 25 million eligible children aged 12-17 years have received two doses of the vaccine.^2^ Quick, accurate, and accessible diagnostic testing for SARS-CoV-2 in pediatric populations is critical to keeping children in classrooms, especially given the transmissibility of newer variants (e.g. delta, omicron) among vaccinated individuals.^3,4^ The impact of disrupted in-person learning has been substantial. Students are more behind in foundational coursework, with the effect compounded for students with historical racial or socioeconomic disadvantages.^5^

One strategy to keep children safely in school is with routine testing. Challenges around testing include delayed result reporting, high cost for reverse transcription polymerase chain reaction (“RT-PCR”) tests, and access disparities among marginalized populations.^6^ Rapid antigen tests offer an attractive alternative with results often returned in 15 minutes, lower cost, and ability to predict patients harboring culturable, infectious virus.^7^ Studies of the accuracy of these tests in children are conflicting. Most were conducted before the more aggressive variants like delta became prevalent.^8,9^ We evaluated the real-world characteristics of the Abbott BinaxNOW rapid antigen test (Abbott Laboratories, Abbott Park, IL) in children presenting to a high-throughput setting. Our study took place in the context of a high prevalence of the delta variant, between May 7, 2021 and December 6, 2021.The prevalence of the delta variant rapidly increased from less than 2.5% of COVID-19 cases to more than 99% in Maryland and surrounding states during the first month of the study period.^10^

## METHODS

### Study design

This single-center prospective study compared the performance, quantified as sensitivity and specificity, of the BinaxNOW rapid antigen test against the current gold standard of RT-PCR. BinaxNOW is a lateral flow assay that targets the SARS-CoV-2 viral nucleocapsid (N) protein. The RT-PCR assay includes a panel of primer/probe sets targeting the viral N gene.

This study was approved by the Johns Hopkins institutional review board.

### Participants and study site

All individuals under 18 years old were offered enrollment from 5/7/2021 – 12/6/2021. Potential enrollees and guardians were given written and verbal information about the study and given the opportunity to opt out of the additional anterior nares swabs for the rapid antigen test. The study site was a high-volume walk-up community collection site linked to the Baltimore Convention Center Field Hospital.

### Data collection

Before sample collection, data for demographics, symptoms, and any self-perceived exposure status was collected for each participant. Presence of active COVID-19 symptoms was assessed based on the standard Centers for Disease Control and Prevention (CDC) symptom checklist.^11^ Participants who reported at least one symptom were considered “symptomatic,” while those who reported no symptoms were considered “asymptomatic.” Participant exposure status was obtained according to CDC risk stratification. Participants were asked about living with someone with confirmed or suspected COVID-19; if they had been within 6 feet of someone with confirmed or suspected COVID-19 for more than 15 minutes; and if they had any other exposure to anyone with confirmed or suspected COVID-19. Participants were considered to have had high risk exposures if they lived with or were within 6 feet for more than 15 minutes of someone with confirmed or suspected COVID-19. Date of symptom onset and exposure were recorded. Test implementation, education, and training processes have been previously described.^12^

### Sample collection

The rapid antigen and RT-PCR samples were collected sequentially for each participant in the study by medical staff who were trained to perform rapid antigen tests.

Manufacturer’s guidelines were followed in obtaining and processing rapid antigen samples.^13^ Staff collected bilateral anterior nares swabs first for direct inoculation onto rapid antigen tests followed by additional bilateral anterior nares sample for RT-PCR. A designated, trained reader interpreted and recorded the result on-site for each rapid antigen test 15 minutes after the test was administered per the instructions for use. All samples for RT-PCR were sent to the University of Maryland Pathology Associates-Maryland Genomics reference laboratory (University of Maryland School of Medicine, Baltimore, MD) and processed using a modification of the CDC 2019 Novel Coronavirus Real-Time RT-PCR Diagnostic Panel. Individuals for whom the RT-PCR or rapid antigen test was deemed indeterminate (control line not interpretable) were excluded from analysis.

### Statistical analysis

Accuracy results (sensitivity, specificity, and positive and negative predictive values [PPV, NPV respectively]) and 95% confidence intervals (CIs) were calculated using the binomial exact method for the rapid antigen test for both symptomatic and asymptomatic populations compared with the RT-PCR gold standard. Accuracy results for symptomatic and asymptomatic group were compared. Two tailed P values were calculated using Fisher’s exact test. Similar test accuracy was calculated for exposure groups and age groups. Cycle threshold (C_T_) values for PCR-positive tests were summarized and compared for symptomatic and asymptomatic groups. A plot showing the distribution of RT-PCR C_T_ values was generated for symptomatic and asymptomatic individuals (Figure 1). All statistical analyses were performed using JMP Pro, version 14.0.0, software (SAS Institute, Cary, NC).

**Figure 1.**
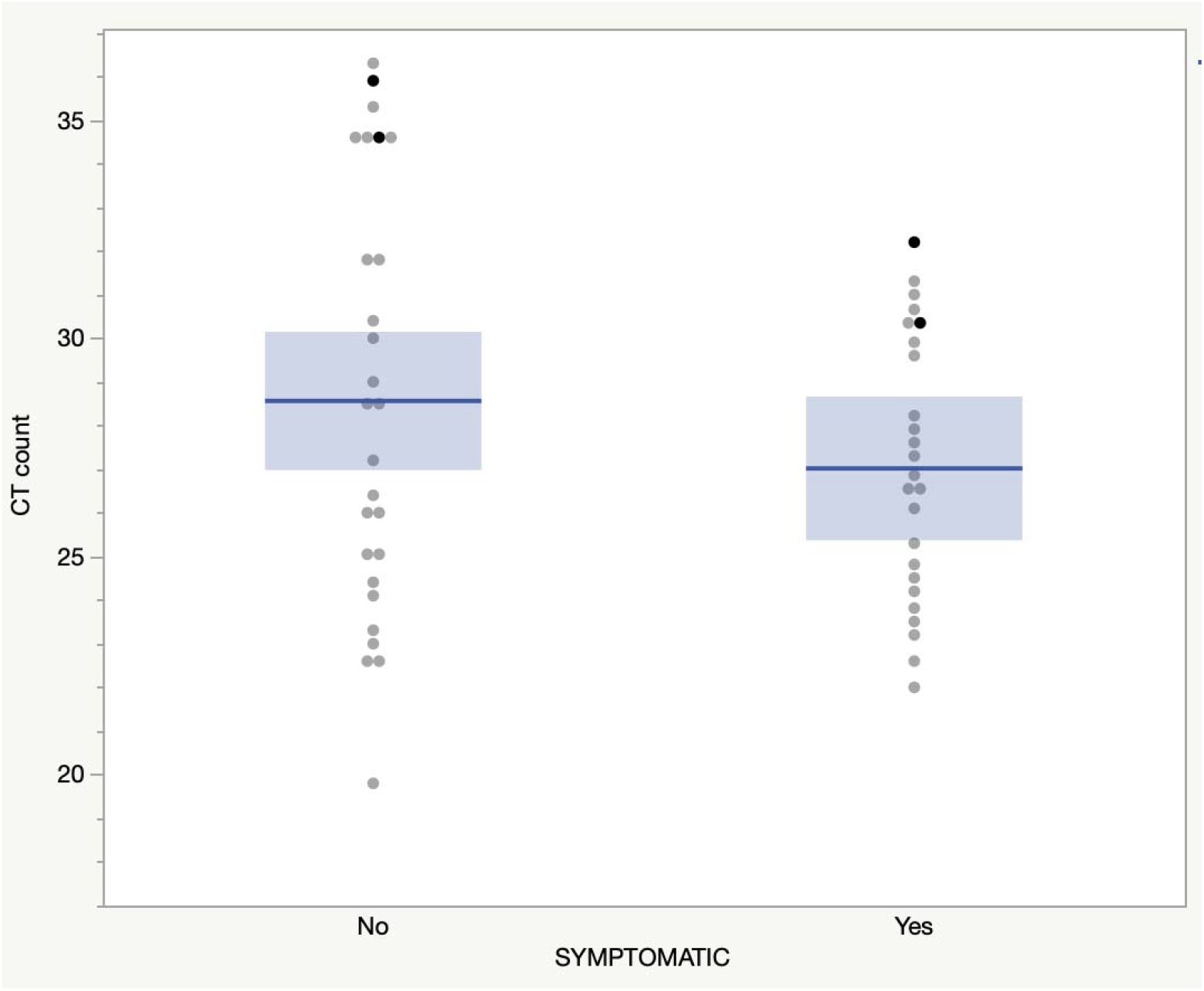
CT counts for symptomatic and asymptomatic participants *Darker dots represent false negative rapid antigen results

## RESULTS

During the testing period, 1054 of 2811 children who were seen for diagnostic testing participated in the study. Among participants, 508 (48.3%) were female, 438 (41.6%) were White, 373 (35.4%) were African American, 105 (9.9%) were Hispanic, and the mean age was 8.9 years. Non-participants were similar for gender (female 49.1%, P = 0.6), were older (mean 9.4 years, P = 0.001), and were more often African American (55%, P < 0.0001) or Hispanic (15.1%, P < 0.0001). Symptomatic status was obtained for 1046 (99.2%) participants, of which 756 (72.4%) were asymptomatic. Symptomatic participants were younger (7.8 vs 9.3 years, P < 0.0001). High risk exposure was reported by 152 (20.1%) asymptomatic and 50 (17.2%) symptomatic participants. The COVID-19 prevalence rate, based on RT-PCR results, was 5.2% (55/1054) overall, 9.0% (26/290) for symptomatic individuals, and 3.6% (27/756) for asymptomatic individuals. For symptomatic participants, 96.2% (275/286) tested within 7 days after symptom onset. The prevalence rate was 20% for symptomatic participants with high-risk exposure and 7.2% for asymptomatic with high-risk exposure (Table 1).

**Table 1.**
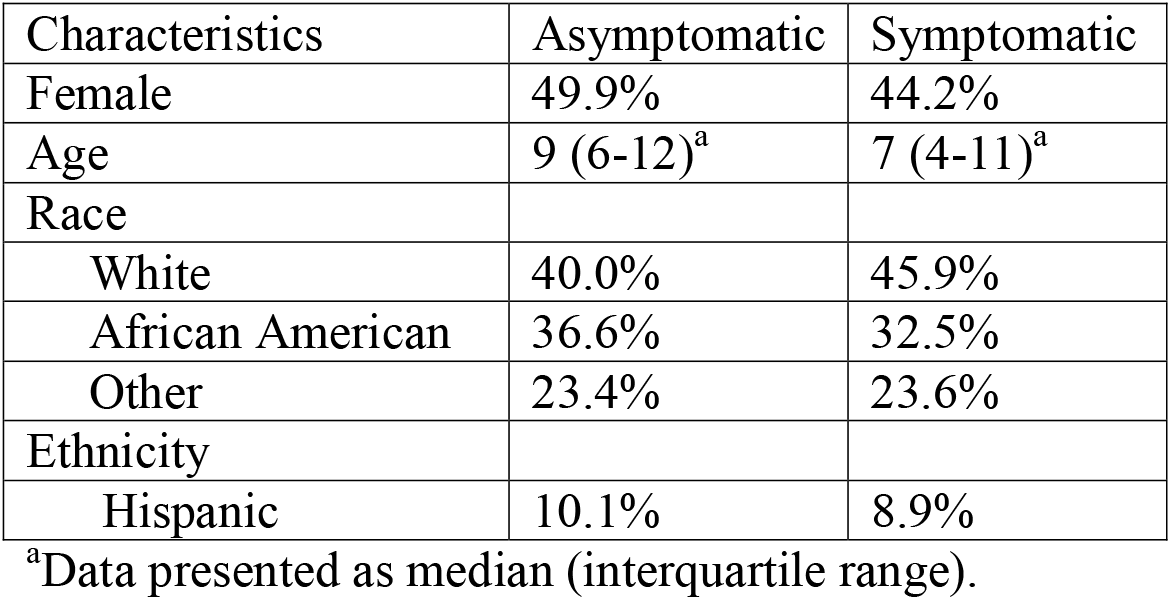
Demographics of Children Presenting for COVID-19 Diagnostic Testing with Concurrent RT-PCR and Rapid Antigen Test, Baltimore, 2021

| Characteristics | Asymptomatic | Symptomatic |
| --- | --- | --- |
| Female | 49.9% | 44.2% |
| Age | 9 (6-12)<sup>a</sup> | 7 (4-11)<sup>a</sup> |
| Race |  |  |
| White | 40.0% | 45.9% |
| African American | 36.6% | 32.5% |
| Other | 23.4% | 23.6% |
| Ethnicity |  |  |
| Hispanic | 10.1% | 8.9% |

<sup>a</sup>Data presented as median (interquartile range).

### Test accuracy

The sensitivity of the rapid antigen tests for all participants was 92.7% (95% CI 82.4% -98.0%) and specificity was 98.0% (95% CI 97.0%-98.8%). Sensitivity was similar for symptomatic participants and asymptomatic participants (92.3% vs. 92.6%) (P = 1.0). It was also similar for high-risk exposure groups, both symptomatic (80.0%; 95% CI 44.4% -97.5%) and asymptomatic (81.8%; 95% CI 48.2% - 97.7%) (Table 2). The specificity for symptomatic participants was 96.6% (95% CI 93.6% - 98.4%) and for asymptomatic participants was 98.6% (95% CI 97.5% - 99.3%). Among symptomatic individuals the PPV was 72.7% (95% CI 54.5% - 86.7%) and the NPV was 99.2% (95% CI 98.2% - 100%).). Among asymptomatic individuals the PPV was 71.4% (95% CI 53.7% - 85.4%) and the NPV was 99.7% (95% CI 99.0% - 100%).

**Table 2.** Antigen Test Accuracy Rates Compared to RT-PCR as Reference Standard

| Exposure Group | Positive/Total (%) |  | Sensitivity<br>N (%) [95%<br>CI] | Specificity<br>N (%) [95%<br>CI] |
| --- | --- | --- | --- | --- |
|  | RT-PCR | Rapid<br>Antigen<br>Test* |  |  |
| Overall | 55/1054<br>(5.2) | 71/1054<br>(6.7) | 51/55 (92.7)<br>[82.4 – 98.0] | 979/999<br>(98.0) [96.9<br>- 98.8] |
| Symptomatic<sup>*</sup><br>- All | 26/290<br>(9.0) | 33/290<br>(11.4) | 24/26 (92.3)<br>[74.9 - 99.1] | 255/264<br>(96.6) [93.6<br>- 98.4] |
| Symptomatic<br>– ≤ 7 days of<br>symptoms | 25/275<br>(9.1) | 32/275<br>(11.6) | 23/25 (92.0)<br>[74.0 - 99.0] | 241/250<br>(96.4) [93.3<br>- 98.3] |
| Symptomatic<br>– High Risk<br>Exposure | 10/50<br>(20) | 10/50<br>(20) | 8/10 (80.0)<br>[44.4 - 97.5] | 38/40 (95.0)<br>[83.1 - 99.4] |
| Asymptomatic<sup>**</sup><br>- All | 27/756<br>(3.6) | 35/756<br>(4.6) | 25/27 (92.6)<br>[75.7 - 99.1] | 719/729<br>(98.6) [97.5<br>- 99.3] |
| Asymptomatic<br>– High Risk<br>Exposure | 11/152<br>(7.2) | 10/152<br>(6.6) | 9/11 (81.8)<br>[48.2 - 97.7] | 140/141<br>(99.3) [96.1<br>– 100.0] |

\*There were no inconclusive or invalid Rapid antigen results \*\* Symptom status data was missing for 8 participants, including 4 participants with rapid antigen positive results RT-PCR= Reverse Transcription Polymerase Chain Reaction CI = Confidence Interval

Mean C_T_ values were similar for the asymptomatic and symptomatic groups (28.6 vs 27) (P = 0.2). The test showed 100% sensitivity at C_T_ count 30 or below in both the symptomatic and asymptomatic populations (Figure 1).

## DISCUSSION

This single center prospective study at a state-owned walk-up testing site showed high sensitivity, specificity, and NPV for the rapid antigen test in both symptomatic and asymptomatic children. The sensitivity was 92.3% in symptomatic participants, and all but one of these 26 RT-PCR positive participants were tested within 7 days of symptom onset. The sensitivity in asymptomatic population was nearly identical at 92.6%, with an NPV of nearly 100%. Cycle threshold counts were similar for symptomatic and asymptomatic individuals and rapid tests showed 100% sensitivity in both these groups at C_T_ count less than 30, which signifies higher viral loads and greater transmissibility. Our point estimate of BinaxNOW rapid antigen test sensitivity in children is above the 80% threshold set by the US Food & Drug Administration (FDA) for Emergency Use Authorization (EUA) for both symptomatic and asymptomatic children.^14^ Our results show that rapid antigen tests provide a reliable means to diagnose and screen for COVID-19 in children. The near 100% NPVs for both symptomatic and asymptomatic children, including some children with recent high-risk exposures, should provide providers and parents with assurance that a negative rapid antigen test with this product can be trusted. These data also provide support to the use of these tests as part of a “test-to-stay” approach in schools.^15^ Because our study showed that roughly 30% of positive antigen tests were false positives, it would be prudent to confirm all positive antigen tests with a PCR test.

Our study shows higher sensitivity and NPV but lower specificity and PPV than other pediatric studies.^8,9^ Differences could be related to higher viral load from prevalent delta variant during the study period as most of the prior pediatric studies were conducted before the delta variant became widespread.^16^ Additionally, ambient conditions for kit storage and use, variation in quality of the test kit between lots, and the skill of our experienced testers may also explain higher sensitivity in our study.^17^

Our study has a few limitations. The results from a single-site with over one-year of experience in high volume testing may not be generalizable to all situations. However, with implementation of best practices, similar accuracy would be expected.^12^ Our study enrolled only a small number of children with high-risk exposure and the estimates of accuracy had broad confidence interval. However, the results are similar to overall symptomatic and asymptomatic groups. Additionally, three of the four false negative tests in the high-risk exposure group were tested within 3 days of exposure and samples were likely collected too early for detection by a rapid antigen test.

Given the short turn-around-time, low cost, and ease of use, this test could play an important role in allowing children to limit absence from school and other activities while in quarantine or awaiting PCR test results especially for asymptomatic children. It may assist in implementing test-to-stay strategies, where exposed school children are allowed to continue uninterrupted in-person learning given frequent testing for one week after exposure.^18^ The higher accuracy reported in our study also underscores the magnitude of missed opportunity in failing to make this important tool widely available to schools. If further studies with the extremely transmissible omicron variant show very high viral load and rapid antigen accuracy, these tests could become one of most valuable tools used to fight current and future COVID-19 variants.

## Data Availability

All data produced in the present work are available upon reasonable request to the authors

